# Electrochemical-biosensor Microchip Based on Gold Nanoparticles as a Point-of-Care Test (POCT) for Quantitative Determination of Glycated Hemoglobin (HbA1c)

**DOI:** 10.1101/2022.10.18.22281233

**Authors:** Kanyarat Boonprasert, Thipaporn Tharavanij, Chiravoot Pechyen, Khanittha Ponsanti, Benchamaporn Tangnorawich, Vithoon Viyanant, Kesara Na-Bangchang

## Abstract

Monitoring the level of glycated hemoglobin (HbA1c) has become the gold standard measure of diabetes mellitus diagnosis and control in conjunction with FBG and oral glucose tolerance test. The study aimed to investigate the applicability of the newly developed nanoparticle-based electrochemical biosensor – multiwalled nanotubes corporated with gold nanoparticles (POCT-HbA1cMWCNTs/AuNPs) as a routine POCT for detection of HbA1c for the diagnosis of diabetes mellitus (DM). Finger-prick and venous blood samples were collected 108 DM and 98 non-DM subjects for determination of HbA1c and total hemoglobin by POCT-HbA1cMWCNTs/AuNPs in comparison with standard HPLC method. The performance of the POCT-HbA1cMWCNTs/AuNPs was evaluated using the standard cut-off HbA1c level of >6.5%. The sensitivity, specificity, positive predictive value, and negative predictive value of the test were 100.00%, 90.32%, 87.23%, and 100.00%, respectively. The probability of DM diagnosis in a subject with HbA1c >6.5 (positive predictive value) was 87.23% (82/94). The accuracy of the POCT-HbA1cMWCNTs/AuNPs was 94.18%, with %DMV (deviation of the mean value) of 0.25%. The results indicate satisfactory assay performance and applicability of the POCT-HbA1cMWCNTs/AuNPs for diagnosis of DM using the cut-off criteria of HbA1c >6.5.

## Introduction

Diabetes mellitus (DM) is a chronic metabolic disorder categorized by elevated levels of plasma glucose due to defects in insulin secretion (type I), insulin action (type II), or both. A series of metabolic diseases follows, which may lead to permanent and irreparable damage, including amputation, retinopathy, neuropathy, and cardiovascular diseases [1]. The growing prevalence of DM is a global concern. Approximately 541 million people are at increased risk for developing type 2 DM. About half of the patients are unaware of their state of illness, and this may lead to delay in proper diagnosis and management [2]. While there is no definitive remedy for DM, disease progression and complications could be minimized by rapid diagnosis and closely monitoring their blood glucose levels [3]. However, fasting plasma glucose concentrations only provide a snapshot of blood glucose at one point in time of blood sample collection and do not provide a long-term vision for disease management [4]. In addition, the level is affected by many different factors and it is not the most effective biomarker for DM diagnosis and control [5]. Monitoring the level of glycated hemoglobin (HbA1c) has become the gold standard measure of DM diagnosis and control in conjunction with FBG and oral glucose tolerance test [6]. HbA1c is hemoglobin HbA glucose, which is a typical glycosylated protein in the body that has nonenzymatic covalent attachment of glucose to the *N*-terminal valine of the beta-globin chain [7]. Its concentration depends on the erythrocyte lifespan and the blood glucose level. The term HbA1c refers to the ratio of HbA1c to total hemoglobin (Hb) concentration, and the normal range of HbA1c is 5 to 20% of total Hb. The cut-off value of HbA1c ≥6.5% is recommended for the diagnosis of DM according to American Diabetes Association and World Health Organization guidelines [8]. Pre-DM is defined as HbA1c of 5.7-6.4%, and non-DM is defined as HbA1c of 4.0-5.6% [9, 10]. Furthermore, HbA1c is also a useful biomarker of long-term glycemia and a good predictor of lipid profile. Monitoring glycemic control could, thus, benefits DM patients with the risk of cardiovascular disease [11]. Accurate and precise methods for HbA1c measurement are therefore required for better glycemic control.

Currently, there are various methods available for measurement of HbA1c levels in the blood [12]. These methods are based on separation according to ionic charge (ion-exchange chromatography, electrophoresis) [13, 14], affinity binding (affinity chromatography for HbA1c) [15], enzymatic assays [16], colorimetric assays [17], or immunoassays [18, 19]. HPLC is considered as one of the gold standard for HbA1c analysis due to its accuracy in simultaneous quantification of both HbA1c and Hb concentrations in whole blood to obtain the HbA1c percentage [20, 21]. Nevertheless, these methods have limitations such as complexity, availability, requirement for experienced and skilled personnel, cost, time consuming, and interference from endogenous and exogenous compounds [22, 23]. Point-of-care testing (POCT) of HbA1c appears to be a user-friendly approach to timely and accurately diagnose DM compared to the standard method. A number of biosensor-based POCTs for the quantitative determination of HbA1c concentration in blood have been developed. Biosensor for HbA1c detection offers potential analytical and testing devise design with regard to test sensitivity, cost, simplicity, robustness and miniaturization [24, 25]. Among these, electrochemical biosensors respond to biological samples rapidly and are suitable for miniaturization ability, rapid and on-site detection, portability, and flexibility [26].

In the present study, the applicability of the newly developed nanoparticle-based electrochemical biosensor – multiwalled nanotubes corporated with gold nanoparticles (POCT-HbA1c^MWCNTs/AuNPs^) as a routine POCT for detection of HbA1c for the diagnosis purpose was investigated [27]. Green synthesis (green chemistry), instead of the conventional approach using toxic chemicals, was applied for the preparation of the gold nanoparticles. The performance of the test was validated in blood samples obtained from patients with confirmed DM and non-DM subjects.

## Materials and Methods

### Electrochemical-Biosensor Microchip

The development of electrochemical-biosensor microchip based on multi-walled carbon nanotubes/gold-nanoparticles (MWCNTs/AuNPs) modified screen-printed carbon electrode (SPCE) for measurement of HbA1c, and total Hb in blood samples has previously been described in detail [27]. In brief, sensor based on the gold nanoparticles (AuNPs) was synthesized from passion fruit (*Passiflora edulis*) peel extracts. The prepared AuNPs (2 ml) were mixed with multiwalled carbon nanotubes (MWCNTs, 5 mg) in various ratios (1:1, 1:2, and 2:1) to form MWCNTs/AuNPs composites. The prepared MWCNTs/AuNPs were characterized for morphology (Transmission electron microscope: TEM), particle size (Dynamic light scattering), crystallinity (X-ray diffraction), functional groups (Fourier transform infrared spectra), UV-absorbance (UV-vis-spectrophotometry), and elemental composition (Energy-dispersive X-ray spectrometry). The nanoparticles are spherical with a diameter of approximately 18 nm, with the crystal structure of face-centred cubic and maximum UV-vis peak at 550 nm. The small AuNPs and the intermeshing of the MWCNTs improve conductivity of composite material. This property, in conjunction with the carbon nanotubes result in good stability and good supports on the fabrication of an electrochemical sensing system. The MWCNTs/AuNPs composite was drop-casted on the SPCE for MWCNTs/AuNPs/SPCE working electrode preparation. The working electrode was air-dried to remove all solvents. The electrochemical response of modified electrodes to WMCNTs:AuNPs (1:1, 1:2, 2:1) was measured using cyclic voltammetry (CV) by PalmSens4 Potentiostat/Galvanostat/Impedance Analyzer (PalmSens, Netherlands). The electrochemical cells consisted of a MWCNTs/AuNPs electrode (working electrode), Ag/AgCl (reference electrode), and platinum electrode (counter electrode). The format of POCT-HbA1c^MWCNTs/AuNPs^ is shown in Fig 1.

**Fig 1.**
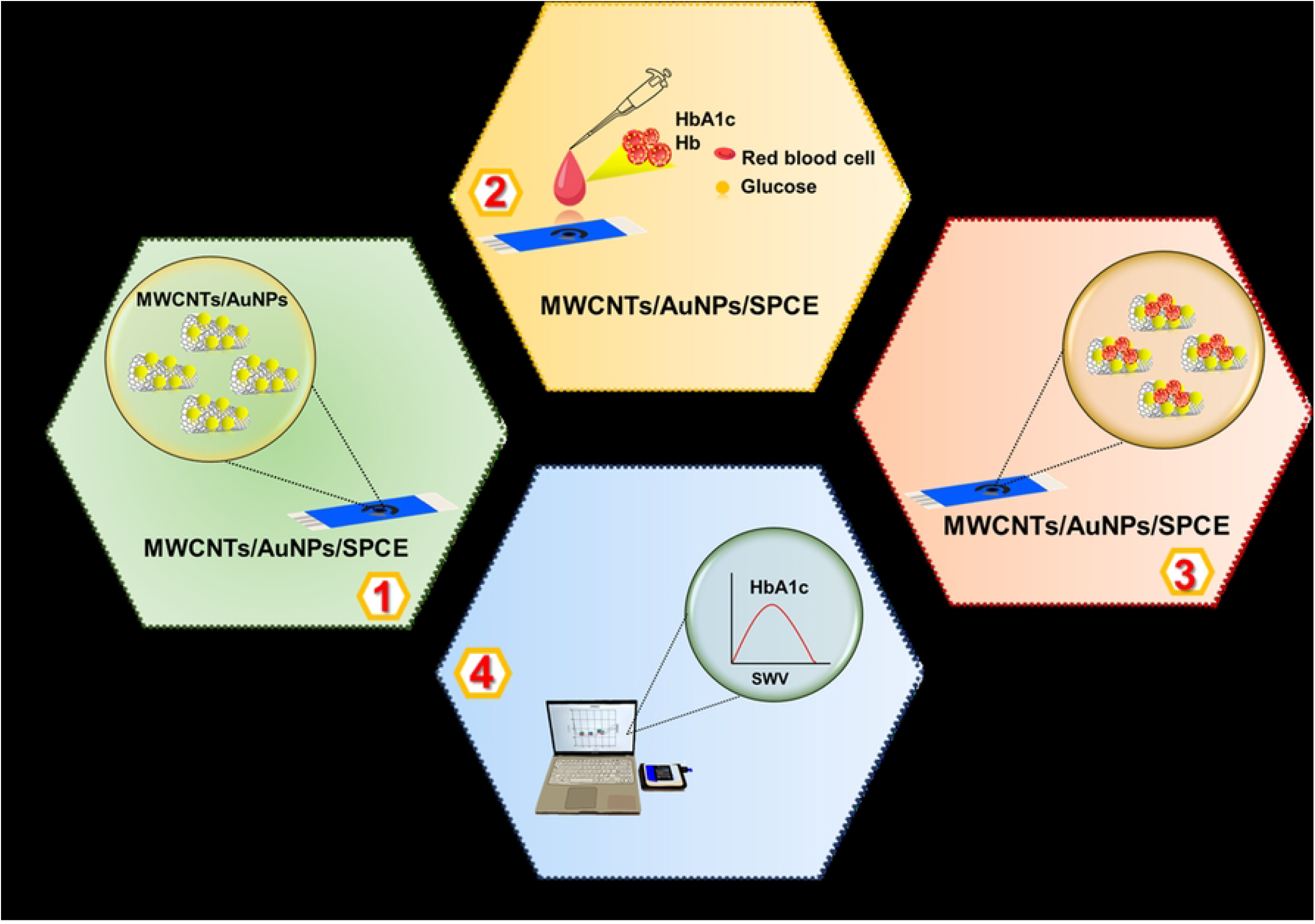
The format of POCT-HbA1c ^MWCNTs/AuNPs^ (Adapted from ref.27).

The responses of MWCNTs/AuNPs/SPCE electrodes to different concentrations of total Hb and HbA1c were investigated by square wave voltammetry (SWV) over the concentration range 5-13 g/dl and 0.186-2.044 g/dl, respectively to construct the calibration curves. The percentage of HbA1c was calculated according to DCCT/NGSP: HbA1c (%) = (HbA1c/Hb)x91.5+2.15 [28, 29].

### Patients

The study was approved by the Ethics Committee, Faculty of Medicine, Thammasat University (approval number 157/2564, project number MTU-EC-00-4-062/64). The study was conducted at Thammasat Chalermprakiet Hospital and Thammasat University Center of Excellence in Pharmacology and Molecular Biology of Malaria and Cholangiocarcinoma, following the guidelines outlined in the Declaration of Helsinki. Written informed consents were obtained from all subjects before study participation. A minimum of 138 study subjects (69 each for DM and non-DM subjects) were recruited for the study. The sample size was estimated using the formula:

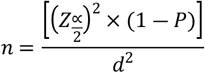

Where n = sample size, Z = 95% confidence interval (1.96), and d (%error) = 5%. P is the sensitivity (0.96) and specificity (0.90) of the POCTs reported in previous studies [30].

Inclusion criteria for the DM group were (i) clinically and laboratory-confirmed DM of all types and stages (ii) males or females (non-pregnant or non-lactating), (iii) aged between 20 and 70 years, and (iv) willing to participate in the study. Exclusion criteria were those with (i) significant abnormality on physical examination, (ii) presence of significant diseases that may affect the study, (iii) abnormal coagulation or concurrent use of anticoagulant/antiplatelet agents, or (vi) participation in any other study within the past 90 days.

Inclusion criteria for the non-DM group were (i) males or females (non-pregnant or non-lactating), (ii) aged between 20 and 70 years, (iii) body mass index (BMI) between 18 and 25 kg/m^2^, (iv) non-smokers and non-alcoholic drinkers, and (vii) willing to participate in the study. Exclusion criteria were those with (i) significant abnormality on physical examination, (ii) history of or having hepatitis B or hepatitis C virus or HIV infection, (v) abnormal coagulation or concurrent use of anticoagulant/antiplatelet agents, or (vi) participation in any other clinical study within the past 90 days.

Blood samples were collected from all volunteers after overnight fasting (8-10 hours). Venous blood samples (3 ml each) were collected in EDTA tubes and transferred to the Bangkok Pathology-Laboratory Co. Ltd. for determination of HbA1c (Automated HbA1c, BioRad D-10 HPLC), total Hb, red blood cell count (RBC), hematocrit, mean cell volume (MCV), mean cell hemoglobin (MCH), mean cell hemoglobin concentration (MCHC), red blood cell distribution width (RDW), platelet count, white blood cell count (WBC). The cut-off value of DM diagnosis is HbA1c ≥6.5%.

### Measurement of HbA1c and Total Hb Concentrations in Blood Samples

Twenty microliters (20 µl) of finger-prick blood were mixed with 50 µl of hemolyzing reagent (consisting of 0.9% tetradecylammonium bromide 26.5 mmol/l), and an aliquot of cell suspension (5 µl) was dropped onto a MWCNTs/AuNPs/SPCE electrode. Electrochemical response was measured using SWV by PalmSens4 Potentiostat/Galvanostat/Impedance Analyzer (PalmSens, Netherlands).

### Evaluation of the performance of POCT-HbA1c ^MWCNTs/AuNPs^

*POCT-HbA1c*^*MWCNTs/AuNPs*^ *performance:* The performance (sensitivity, specificity, positive and negative predictive values, and accuracy) of the POCT-HbA1c^MWCNTs/AuNPs^ was evaluated by comparing the measured HbA1c values with those measured by the reference laboratory method using the standard cut-off HbA1c level of ≥6.5% [9]. The test performances were estimated as follows:

Sensitivity (%) = TP/(TP+FN)

Specificity (%) = TN/(TN+FP)

Positive predictive value (%) = TN/(TN+FN)

Negative predictive value (%) = TP/(TP+FP)

Accuracy (%) = (TP+TN)/(TP+TN+FP+FN)

False-positive (%) = Number of misdiagnosed DM cases by POCT/Total number of negative cases by reference method

False-negative (%) = Number of misdiagnosed non-DM cases by POCT/Total number of positive cases by reference method

Where TP = True positive, TN = True negative, FP = False positive, FN = False negative.

#### Test agreement analysis

The 95% confidence interval and the mean relative error were estimated by the Bland-Altman plot and PasinggeBablok regression analysis (MedCalc Software, Mariakerke, Belgium) to determine the consistency of the HbA1c values measured by POCT-HbA1c^MWCNTs/AuNPs^ and those measured by the reference laboratory method.

### Statistics

The statistical analysis was performed using SPSS 17.0 software. Quantitative variables are summarized as median (range) and mean (SD) values for variables with non-normal and normal distribution, respectively. Deviation of the mean HbA1c value measured by POCT test from the value measured by the reference laboratory method was expressed as %DMV (% deviation from mean value). The correlation between the concentrations of the two quantitative variables was determined using Spearman’s or Pearson’s correlation test for variables with non-normal or normal distribution, respectively. Statistical significance was set at α=0.05.

## Results

### Electrochemical Detection of Total Hb

The responses of MWCNTs/AuNPs/SPCE electrodes (SWV) to different total Hb concentrations are shown in Fig 2. The MWCNTs/AuNPs/SPCE electrodes generated an oxidation peak at -0.3 V. The intensity of the peak current was proportional to the concentration of total Hb over the concentration range of 5-13.8 g/dl. The position of the oxidation peak remained unchanged with the increased concentrations of total Hb. The limit of detection was 5 g/dl. The equation for linear regression was: IP (µA) = 1.228C+21.287, with a correlation coefficient (r) of 0.9004 (Fig 2).

**Fig 2.**
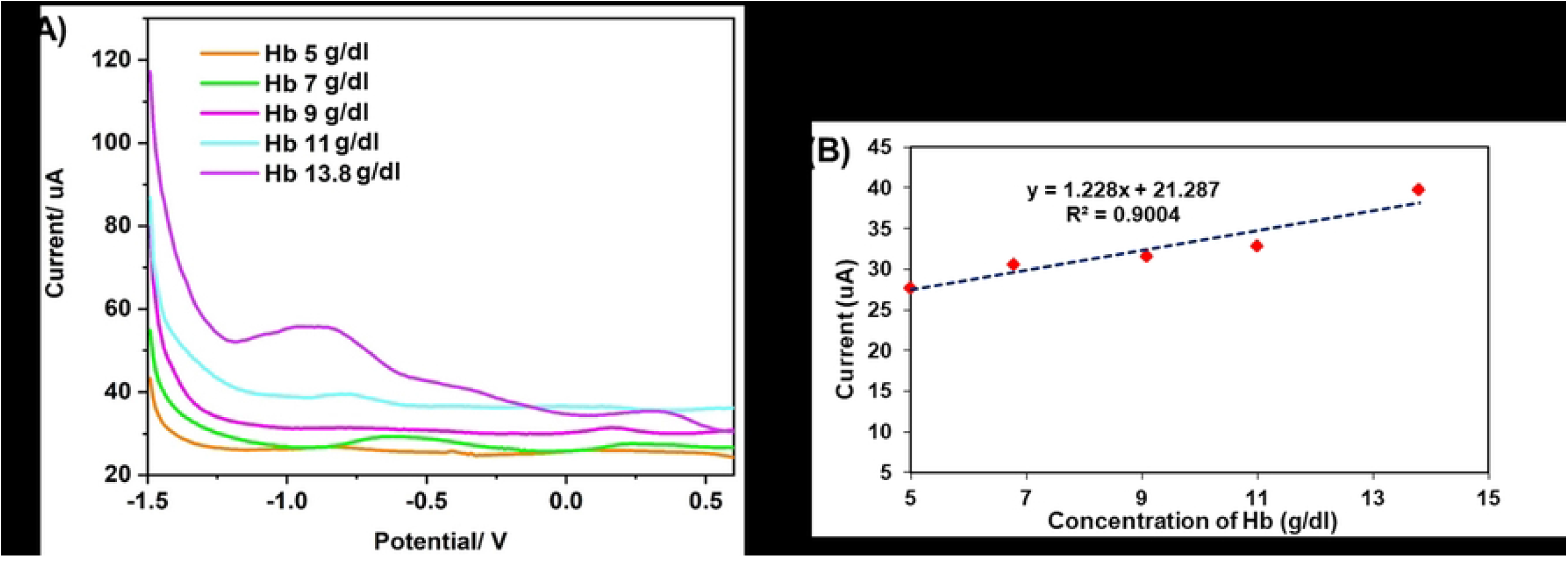
(A) SWV voltammograms for detection of total Hb with MWCNTs/AuNPs/SPCE electrodes, and (B) Plot of current vs total Hb concentration (Adapted from ref.27).

### Electrochemical Detection of HbA1c

For SWV, the peak current was increased with increased Hb1Ac concentrations (0.186-2.044 g/dl) (Fig 3). The relationship between the current and Hb1Ac concentration was linear: I_P_ (µA) = 8.6488C+11.122, r=0.9645). The limit of detection was 0.186 g/dl. The specificity test for HbA1c was carried out against Hb, BSA and glucose under identical experimental conditions.

**Fig 3.**
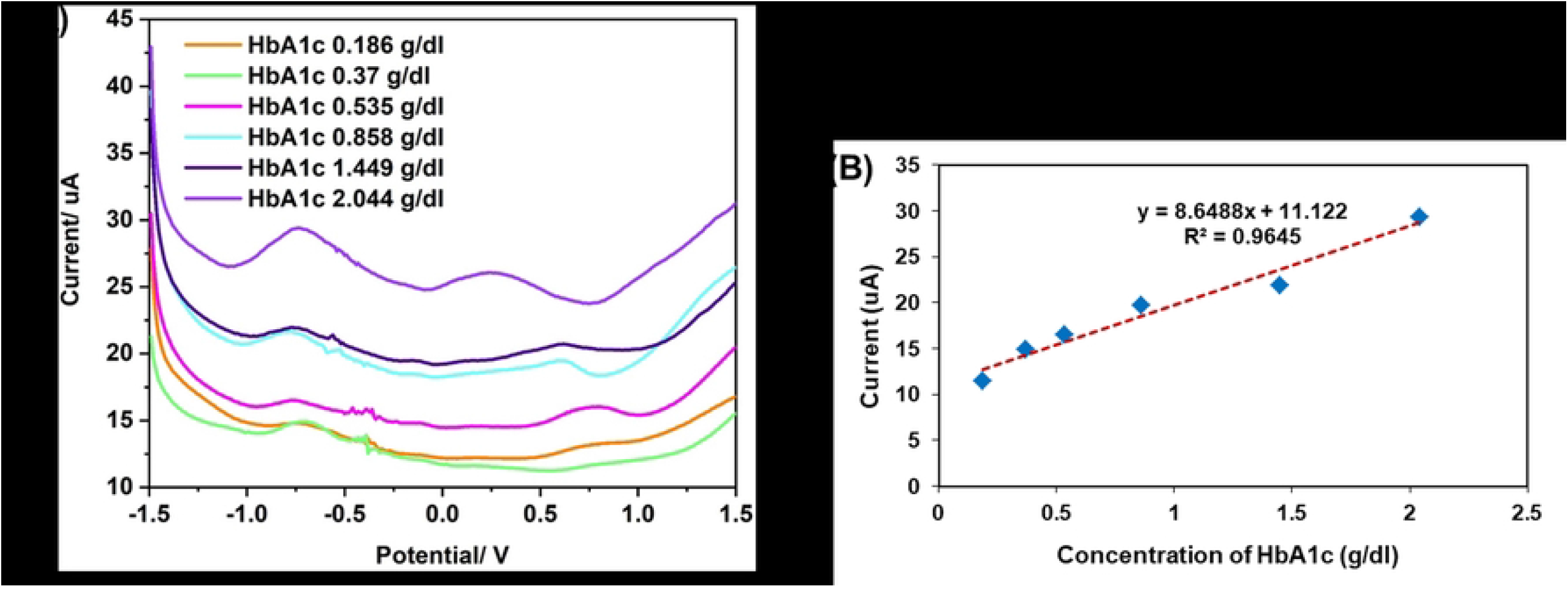
(A) SWV voltammograms for detection of HbA1c with MWCNTs/AuNPs/SPCE electrodes, and (B) Plot of current vs HbA1c concentration (Adapted from ref.27).

### Analysis of the Performance of POCT-HbA1c^MWCNTs/AuNPs^

Analysis of the performance of the POCT-HbA1c compared with the reference laboratory method was performed in 108 DM and 98 non-DM subjects. Median (range) HbA1c concentrations measured by POCT-HbA1c^MWCNTs/AuNPs^ in DM patients were 0.64 (0.36-0.93) and 0.48 (0.22-0.64) g/dl, respectively [corresponding to 7.19 (5.18-11.89) and 6.03 (4.10-6.40) % of total Hb, respectively]. The corresponding HbA1c concentrations measured by the reference method were 0.70 (0.39-1.78) and 0.45 (0.25-0.66) g/dl, respectively [corresponding to 7.10 (4.80-14.00) and 5.30 (4.30-6.40) % of total Hb, respectively). Median (range) total Hb concentrations measured by POCT-HbA1c^MWCNTs/AuNPs^ in DM patients were 12.65 (10.26-14.71) and 11.46 (9.65-12.58) g/dl, respectively. The corresponding total Hb concentrations measured by the reference method were 13.10 (10.50-15.50) and 13.00 (8.80-15.20) g/dl, respectively.

#### POCT-HbA1c^MWCNTs/AuNPs^ performance

To evaluate the clinical applicability of the POCT-HbA1c^MWCNTs/AuNPs^, the HbA1c values measured by the POCT-HbA1c^MWCNTs/AuNPs^ were compared with that measured by the reference laboratory method using the cut-off value of 6.5% (Table 1). The sensitivity and specificity of the POCT-HbA1c were 100.00% (82/82) and 90.32% (112/124), respectively. The probability of DM diagnosis in a subject with HbA1c ≥6.5 (positive predictive value) was 87.23% (82/94). The probability of misdiagnosis in a subject with HbA1c <6.5 (negative predictive value) was 100.00% (112/112). The accuracy of the POCT-HbA1c^MWCNTs/AuNPs^ expressed as a proportion of true positives and true negatives (correctly classified by POCT-HbA1c^MWCNTs/AuNPs^) in all subjects was 94.18%, with %DMV of 0.25%.

**Table 1.** Analysis of HbA1c levels using POCT-HbA1c ^MWCNTs/AuNPs^ based on the 6.5% cut-off value. Data are presented as the number of subjects (n=206).

| HbA1c measured by<br>POCT (%) | HbA1c measured by reference laboratory method (%) |  |
| --- | --- | --- |
| | HbA1c ≥ 6.5 | HbA1c < 6.5 |
| HbA1c ≥ 6.5 | 82 | 12 |
| HbA1c < 6.5 | 0 | 112 |
| **Total** | 82 | 124 |

#### Test agreement analysis

Bland-Altman plot analysis of the HbA1c analyzed by POCT-HbA1c^MWCNTs/AuNPs^ and the reference methods was performed, and results showed that only 12 subjects had values outside the 95% CI (confidence interval) of the range of agreement limit (−1.65614 to 1.36814), suggesting that 94.18% of the value measured by POCT-HbA1c^MWCNTs/AuNPs^ was in agreement with that measured by the standard method (Fig 4).

**Fig 4.**
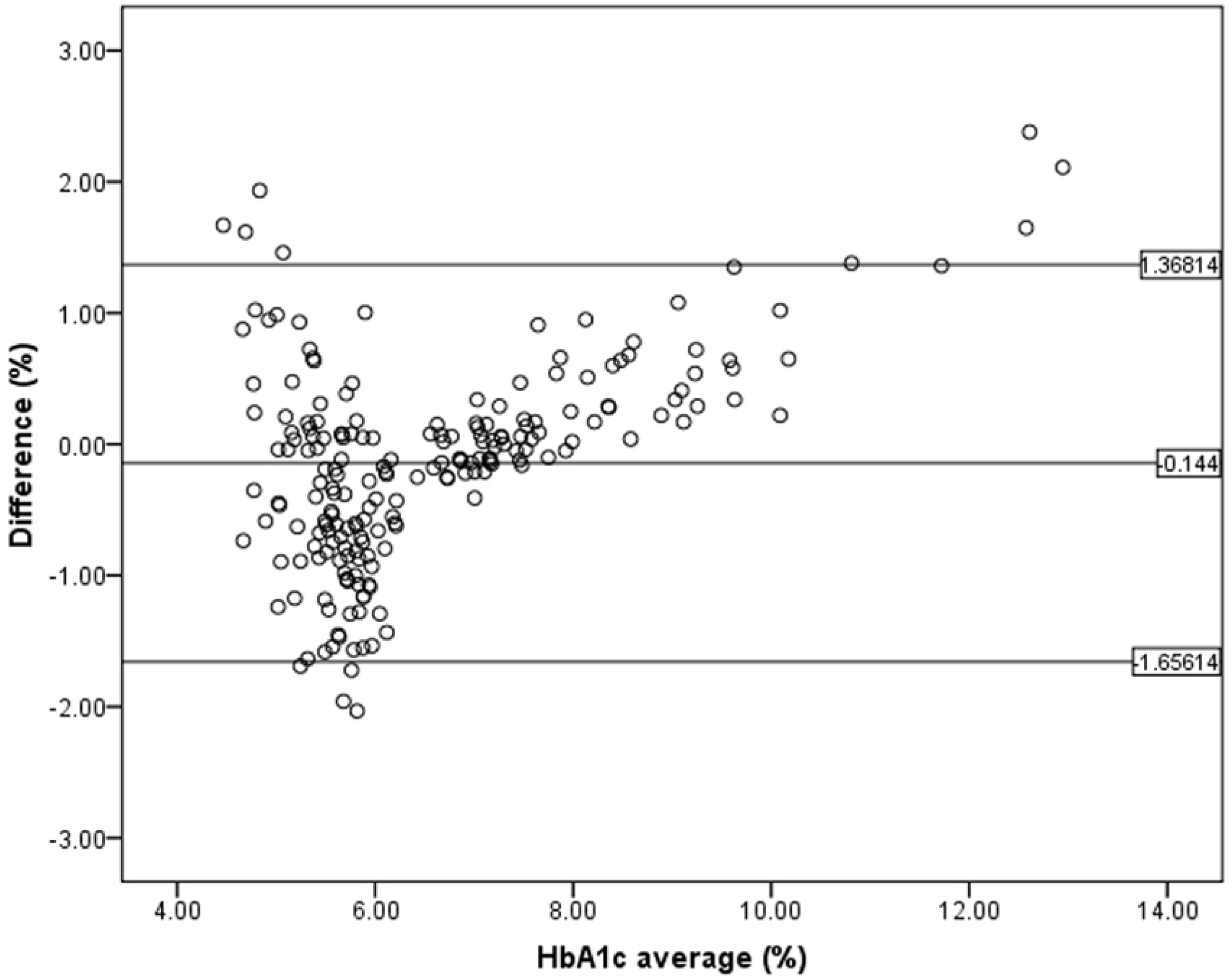
Bland-Altman plot analysis for accuracy of HbA1c measured by POCT-HbA1c compared with the reference method in 206 blood samples collected from DM and non-DM subjects.

### Clinical Application of POCT-HbA1c^MWCNTs/AuNPs^

Clinical application of the POCT-HbA1c^MWCNTs/AuNPs^ was evaluated in 108 DM and 98 non-DM subjects. The demographic and laboratory data of the DM and non-DM subjects are summarized in Table 2. Excellent linear correlation (*p*<0.0001, Y = 2.25+0.69*X, r=+0.874) was found between the HbA1c values measured by POCT-HbA1c and the reference laboratory method (Fig 5). There was a moderate linear correlation between FPG and HbA1c levels measured by the standard methods in DM and non-DM subjects (r=0.552, *p*<0.0001).

**Table 2.**
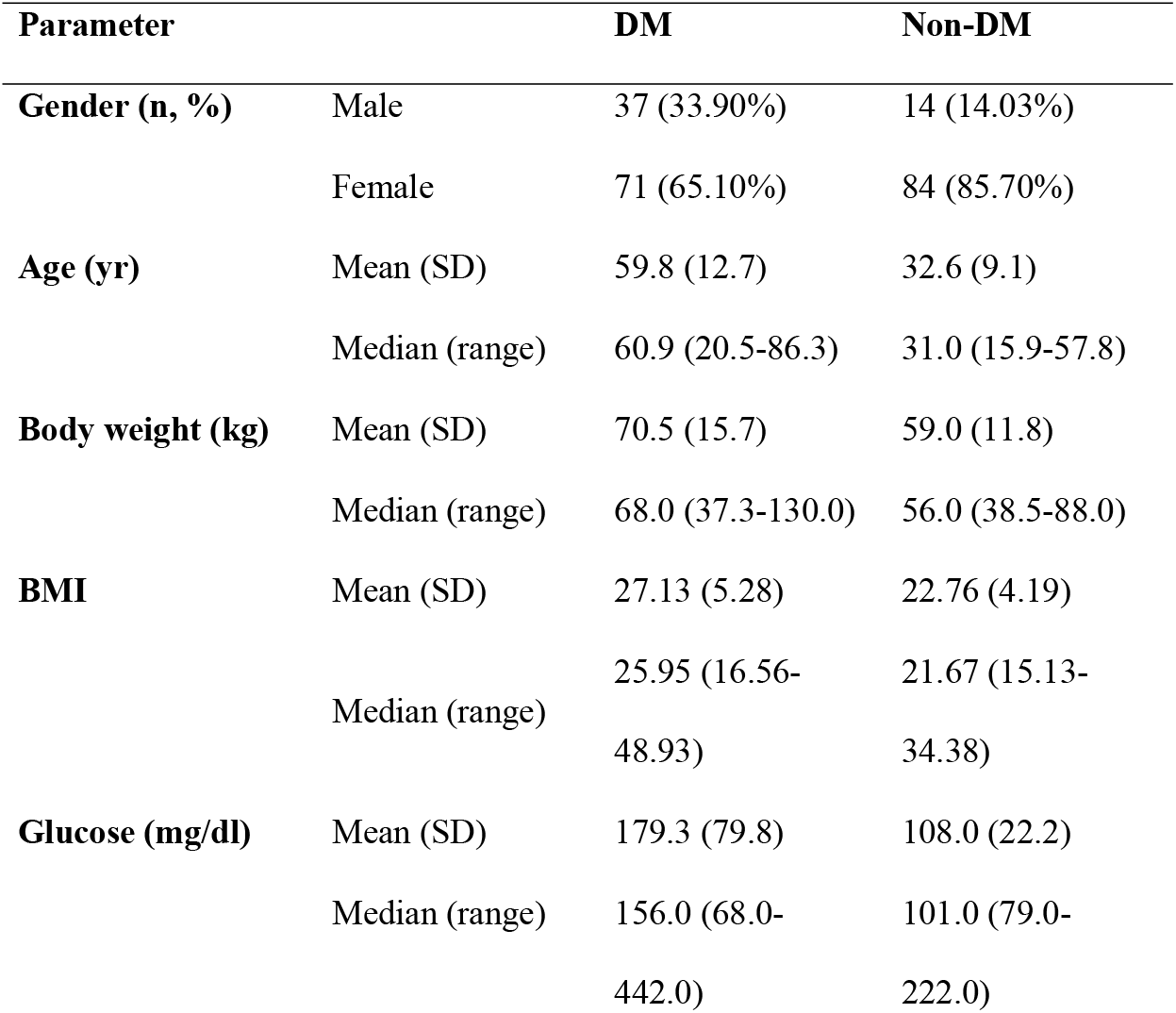
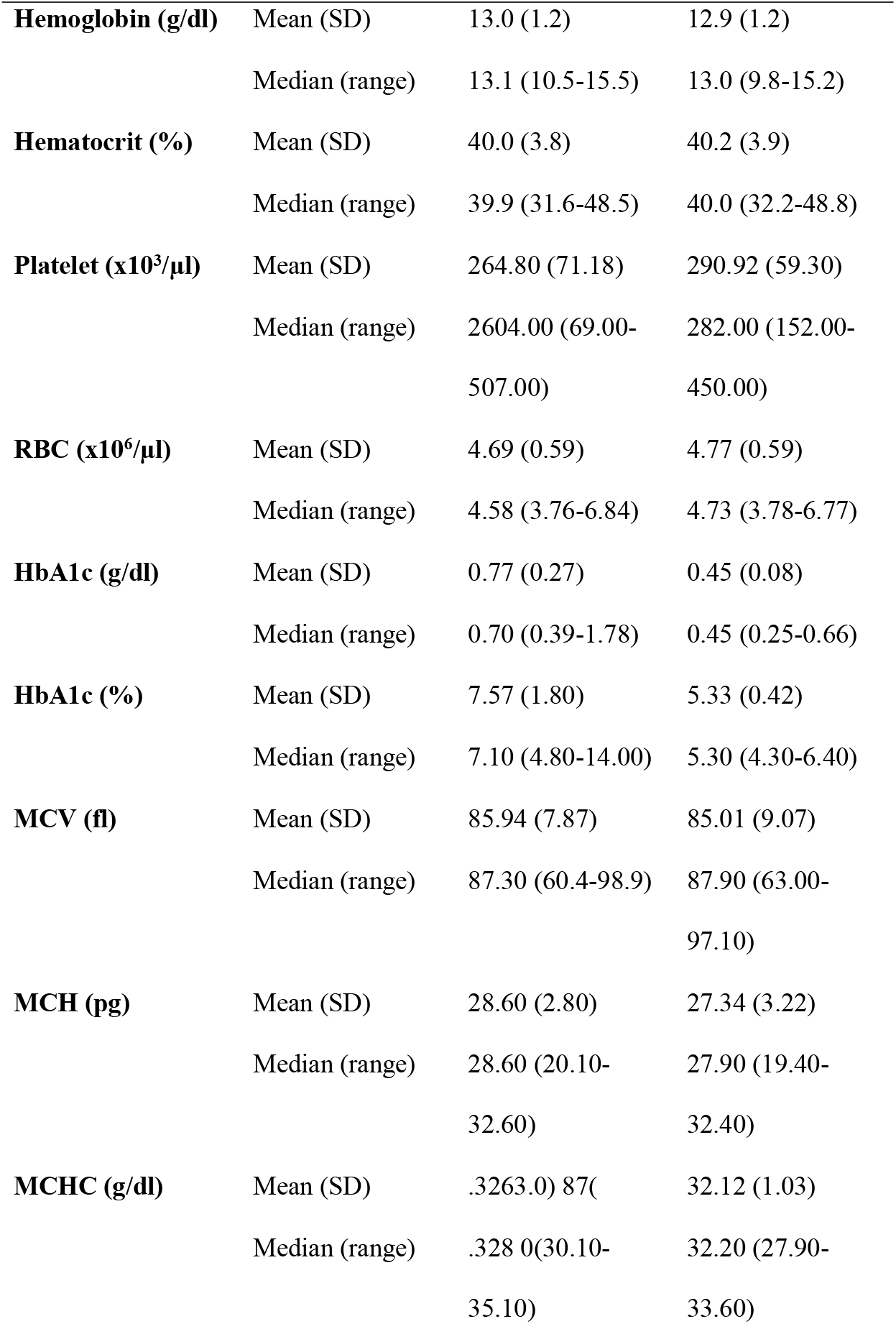
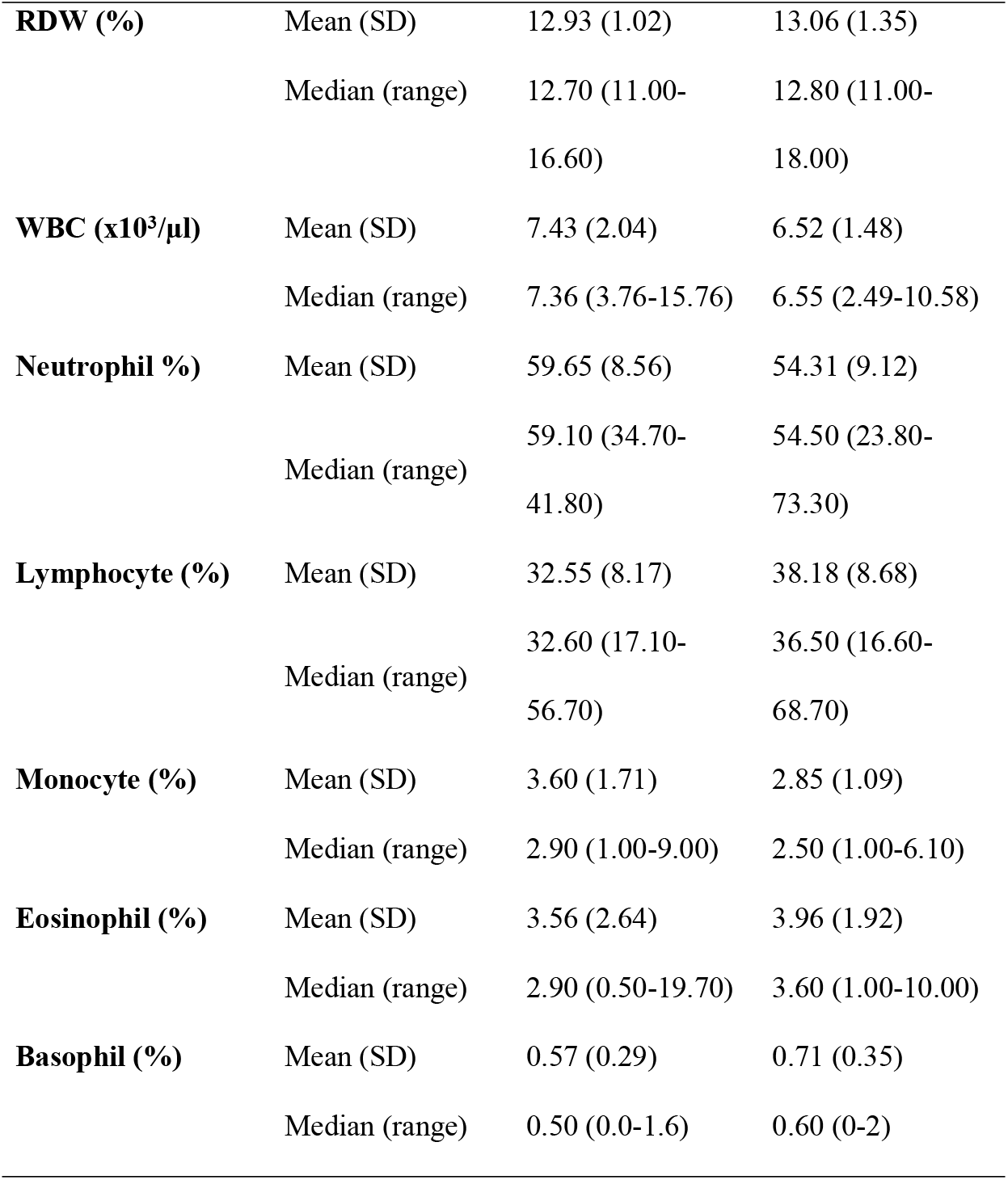
Demographic, glucose level and hematological data of DM and non-DM subjects. Data are presented as number (%) or mean (SD) and median (range) values.

**Fig 5.**
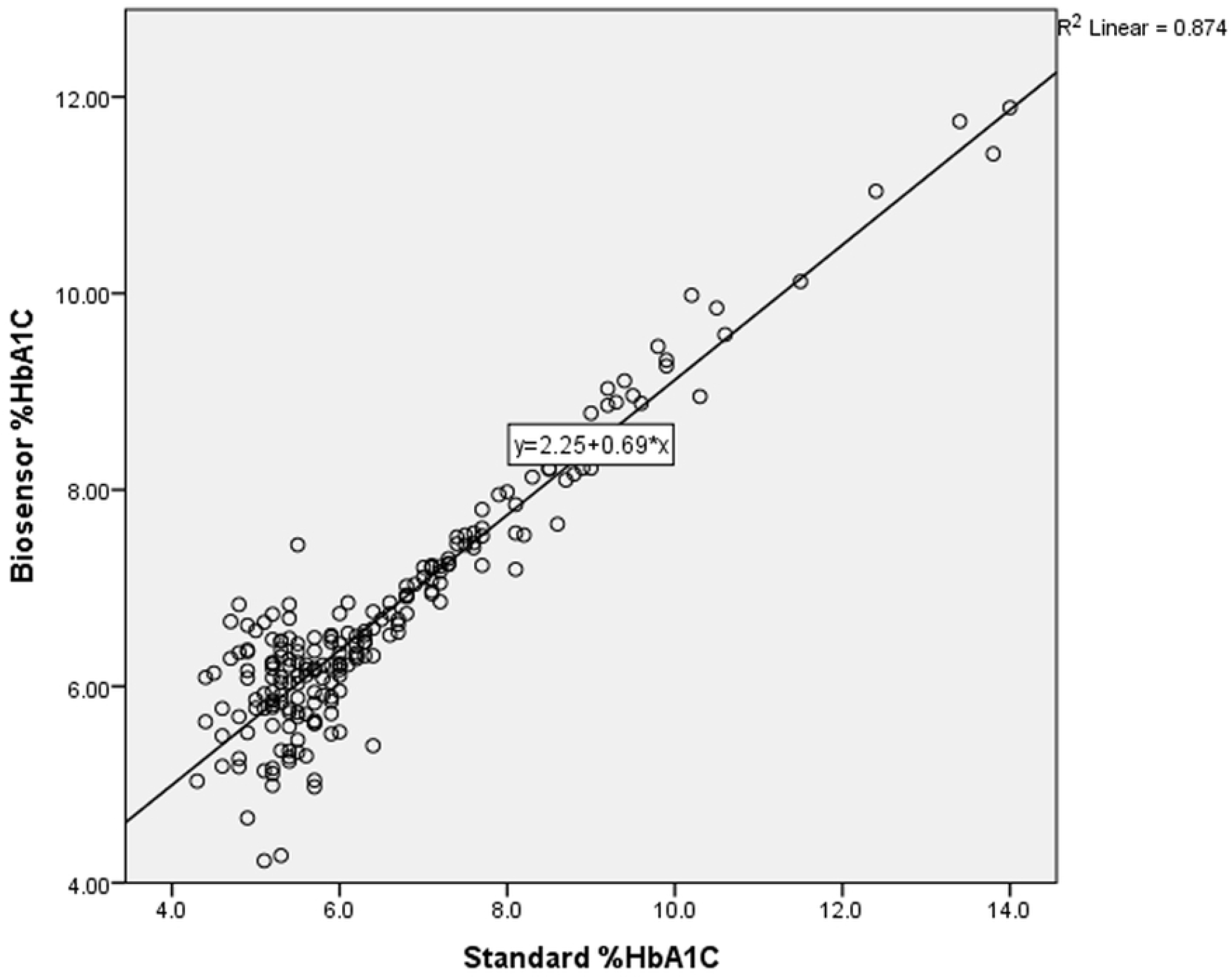
Correlation between HbA1C concentrations measured by POCT-HbA1c ^MWCNTs/AuNPs^ and the reference laboratory methods in 206 subjects (108 DM and 98 non-DM subjects). (*p* < 0.0001, r=+0.874).

Correlation analysis of the HbA1c values measured by POCT-HbA1c^MWCNTs/AuNPs^ and hematological parameters (Hb, Hct, MCV, MCHC, and RDW) was performed to determine hematological factors that could influence the measurement of HbA1c by POCT-HbA1c^MWCNTs/AuNPs^. Only two parameters were found to be significantly correlated with the measured HbA1c by POCT-HbA1c^MWCNTs/AuNPs^. Total Hb concentration (measured by POCT-HbA1c^MWCNTs/AuNPs^ and reference method) was weakly (r=+0.310) but significantly (*p*<0.0001) correlated with the HbA1c measured by POCT-HbA1c^MWCNTs/AuNPs^. On the other hand, a negative correlation (r=-0.214, *p*=0.002) was found between MCHC and HbA1c values measured by POCT-HbA1c^MWCNTs/AuNPs^.

## Discussion

The glycosylated Hb is formed *via* a spontaneous non-enzymatic reaction between glucose and the *N*-terminal residues of beta-globin chain in Hb to generate unstable Schiff base intermediates. Then, the intermediate goes through an irreversible Amadori rearrangement to synthesize the stable ketoamine [31]. The accomplishment of quantitative HbA1c provides in-depth and accurate detection for clinical application. A number of HbA1c POCT devices are currently available in clinical settings, of which the detection method is either affinity-based separation or immunoassay techniques. Biosensor with electrochemical detection has also been developed as POCT for HbA1c. Each has some limitations, *e*.*g*., test complications requiring multi-steps of sample preparation [32], and the requirement of large blood sample volume [33]. Furthermore, most are subject to interferences from Hb variants (HbC, HbE, HbS, and others) and modified Hb (carbamylated and acetylated Hb, or labile HbA1c) [34]. The analysis showed that most of the HbA1c POCT devices have a mean negativity bias compared to laboratory assays, as well as a high variability among the bias value within devices. Recently, gold nanoflowers modified electrochemical biosensor based on 4-mercaptophenylboronic acid was developed for the quantitative determination of HbA1c [35]. In this study, we investigated the clinical applicability of the POCT-HbA1c ^MWCNTs/AuNPs^ previously developed in our laboratory [27] for the accurate diagnosis of DM. The test was sensitive and specific for the determination of HbA1c in the blood (from both venipuncture and finger-prick), exploiting the catalytic property of HbA1c for the reduction reaction of H_2_O_2_ to produce the electrochemical signal. The biosensor was prepared from gold nanoparticles (AuNPs) in multiwalled carbon nanotubes (MWCNTs) as MWCNTs/AuNPs composites. The obvious advantage of the test is the use of green synthesis (green chemistry) for the synthesis of AuNPs from passion fruit peel instead of toxic chemicals. In addition, it is the label-free method that assures rapid detection. AuNPs were electrochemically deposited onto the SPCE. The starch composition in the passion fruit peel with different amylose pectins acts as a reducing agent in the electrochemical reaction [36]. This label-free method could assure rapid detection with as small a volume as 20 μl of blood. Only a single step pre-treatment of blood samples is required to remove plasma interference, followed by lysis of the RBC. The POCT-HbA1c^MWCNTs/AuNPs^ provides simultaneous determination of both HbA1c and total Hb concentrations in a single reaction. This feature not only reduces the testing time but also reduces the errors from the use of different reactions.

To demonstrate the clinical applicability of the developed POCT-HbA1c^MWCNTs/AuNPs^ electrochemical biosensor, the concentrations of HbA1c in blood samples from DM and non-DM subjects measured by the POCT-HbA1c^MWCNTs/AuNPs^ and the reference laboratory methods were compared. Results show high sensitivity (100%), specificity (90.32%), accuracy (94.18%), and excellent application prospects in clinical practice. The probability of a diagnosis of DM in subjects with HbA1c level ≥6.5% (positive predictive value) was 87.23%, while the probability of misdiagnosis of DM in subjects with HbA1c level <6.5% (negative predictive value) was 100.00%. The agreement of the values measured by both methods was excellent (r=+0.874). The Bland-Altman plot suggested that 94.18% of the value measured by POCT-HbA1c^MWCNTs/AuNPs^ was in agreement with that measured by the standard method. The HbA1c level was correlated with FPG as previously reported [11]. The measurement of HbA1c as a percentage of total Hb relies on the accuracy of measurement of both HbA1c and total Hb concentrations. A weak linear correlation was found between total Hb concentration (measured by standard and POCT-HbA1c^MWCNTs/AuNPs^ methods) and HbA1c concentration measured by POCT-HbA1c^MWCNTs/AuNPs^ (r=+0.32). This suggests little influence of the total Hb on HbA1c concentration measured by POCT-HbA1c^MWCNTs/AuNPs,^ and the POCT-HbA1c^MWCNTs/AuNPs^ could have wide clinical applicability in patients regardless of the Hb concentration. The influence of total Hb concentration on HbA1c determination has been reported in patients with hemoglobinopathies such as thalassemia. The hematocrit varies among individuals, which leads to differences in total Hb. The low level of hematocrit could be due to the following reasons: the amounts of RBC and Hb in RBC are not enough to completely fill the surface of the sensor, and/or HbA1c does not reflect an actual ratio. On the other hand, the MCHC value, which relates to the volume of RBC, affected the HbAc1 level measured by POCT-HbA1c^MWCNTs/AuNPs^ in a reverse direction; the higher the MCHC concentration, the lower the HbA1c concentration measured by POCT-HbA1c^MWCNTs/AuNPs^ (r=-0.214, *p*=0.002). Falsely high HbA1c level has been reported with conditions that prolong the life-span of RBC (*e*.*g*., iron deficiency anemia), severe hypertriglyceremia (>1,750 mg/dl), severe albuminemia (>20 mg/dl), alcohol intake and administration of some drugs (*e*.*g*., salicylate, opioids) [37]. On the other hand, falsely low HbA1c level has been reported in conditions that shorten the life-span of RBCs and splenomegaly, pregnancy, and administration of some drugs (*e*.*g*., ribavirin, cephalosporin, and levofloxacin).

In conclusion, the results of the study indicate satisfactory assay performance and applicability of the POCT-HbA1c^MWCNTs/AuNPs^ based on gold nanoparticles modified screen-printed carbon electrode for diagnosis of DM using the cut-off criteria of HbA1c ≥6.5%. The POCT-HbA1c^MWCNTs/AuNPs^ is an accurate and easy-to-use tool that provides the test results on the spot and become an effective tool in establishing the DM diagnosis, especially in vulnerable or hard-to-reach populations. It does not require fasting before measurement, and the sample can be stored at 4 C° for at least two weeks in the case when measurement cannot be performed immediately [38, 39]. The user-friendly format in smartphone application (MyA1c) is being developed for clinical use in routine diagnosis of DM. The implication of using POCT-HbA1c^MWCNTs/AuNPs^ in medical treatment decision-making to control DM patients and patient outcomes needs to be further evaluated.

## Data Availability

All relevant data are within the manuscript and its Supporting Information files.

## Abbreviations

(HbA1c): Glycated hemoglobin
(POCT): Point-of-Care Test
(DM): Diabetes mellitus
(POCT-HbA1c^MWCNTs/AuNPs^): Nanoparticle-based electrochemical biosensor – multiwalled nanotubes corporated with gold nanoparticles

## Acknowledgments

We thank Dr. Panida Kongjam (Clinical Coordination, WHO collaborating Center, Thammasat University) for statistical analysis and Miss Akkaracha Hanwattanakul (Faculty of Science and Technology, Thammasat University) for data collection.

## Author contributions

Conceptualization: Kesara Na-Bangchang, Kanyarat Boonprasert, Chiravoot Pechyen

Data Curation: Kesara Na-Bangchang, Kanyarat Boonprasert, Thipaporn Tharavanij

Formal Analysis: Kesara Na-Bangchang, Kanyarat Boonprasert

Funding Acquisition: Kesara Na-Bangchang, Kanyarat Boonprasert

Investigation: Kanyarat Boonprasert, Khanittha Ponsanti

Methodology: Kesara Na-Bangchang, Kanyarat Boonprasert, Chiravoot Pechyen, Benchamaporn Tangnorawich

Project Administration: Kesara Na-Bangchang, Kanyarat Boonprasert

Resources: Kesara Na-Bangchang, Chiravoot Pechyen, Thipaporn Tharavanij

Software: Kanyarat Boonprasert, Chiravoot Pechyen, Khanittha Ponsanti, Benchamaporn Tangnorawich

Supervision: Kesara Na-Bangchang, Thipaporn Tharavanij, Vithoon Viyanant, Benchamaporn Tangnorawich

Validation: Kesara Na-Bangchang, Kanyarat Boonprasert, Vithoon Viyanant

Visualization: Kanyarat Boonprasert, Chiravoot Pechyen, Khanittha Ponsanti

Writing – Original Draft Preparation: Kesara Na-Bangchang, Kanyarat Boonprasert,

Writing – Review & Editing: Kanyarat Boonprasert, Thipaporn Tharavanij, Chiravoot Pechyen, Khanittha Ponsanti, Benchamaporn Tangnorawich, Vithoon Viyanant, Kesara Na-Bangchang

## Declaration of Conflict Interest

The authors declare that they have no known competing financial interests or personal relationships or affiliations that could have appeared to influence the work reported in this paper.

## Funding

The study was supported by the National Research Council of Thailand (Genetic markers and Point-of-care test for Healthy Aging: 169/2563), and a Ph.D. scholarship from Thammasat University.

## Notes

### Competing Interest Statement

The authors have declared no competing interest.

### Funding Statement

No. The funders had no role in study design and analysis, decision to publish, or preparation of the manuscript.

### Author Declarations

The Ethics Committee, Faculty of Medicine, Thammasat University (approval number 157/2564, project number MTU-EC-00-4-062/64).

